# Study protocol for a multicenter randomized controlled trial comparing standby versus prophylactic extracorporeal membrane oxygenation in high-risk percutaneous coronary intervention (ECMO-READY trial)

**DOI:** 10.64898/2026.07.24.26358881

**Authors:** Liangshan Wang, Yan Wang, Andong Lu, Kexin Wang, Chenglong Li, Fang Liu, Xiaolin Yu, Dingyu Wang, Liwen Lyu, Wenlong Duan, Xinguang Wei, Eddy Fan, Zhongtao Du, Hong Wang, Yan Liu, Jianchao Li, Xiaotong Hou, the ECMO-READY investigators

## Abstract

**Background:** Although prophylactic veno-arterial extracorporeal membrane oxygenation (VA-ECMO) may provide hemodynamic stability during high-risk percutaneous coronary intervention (PCI), it is also associated with potential complications and may not be necessary in most cases. In this context, we proposed a pre-cannulated standby ECMO strategy and designed the ECMO-READY trial to evaluate the comparative effectiveness of pre-cannulated standby versus prophylactic ECMO strategies in patients undergoing high-risk PCI.

**Methods:** The ECMO-READY trial is a prospective, multicenter, open-label, randomized controlled trial conducted in 8 sites in China. A total of 176 patients scheduled to undergo high-risk PCI will be randomly assigned in a 1:1 ratio to either a pre-cannulated standby ECMO strategy or a prophylactic ECMO strategy. The primary outcome is the 30-day incidence of major adverse events, including death, myocardial, infarction, repeat revascularization, stroke, PCI failure, limb ischemia, major bleeding, vascular injury requiring intervention, and need for renal replacement therapy. Secondary outcomes include post-procedural hemoglobin decline, post-procedural platelet count decline, red blood cell transfusion rate, peak post-procedural interleukin-6 level, use of intra-aortic balloon pump, duration of ECMO support, length of hospital stay, hospitalization cost, and each component of the composite primary outcome. Enrollment began in March 2025 and is anticipated to be completed by December 2026.

**Discussion:** The ECMO-READY trial will provide prospective randomized evidence regarding ECMO support strategies in patients undergoing high-risk PCI and may help inform future clinical practice.

**Trial registration:** ClinicalTrials.gov NCT06274411. Registered on February 23, 2024.

## Introduction

### Background and rationale {6a}

Over the past 2 decades, percutaneous coronary intervention (PCI) has been increased performed in patients with multivessel or unprotected left main coronary artery disease and impaired left ventricular systolic function[1]. However, in these high-risk patients, transient interruption of coronary blood flow during PCI may cause hemodynamic collapse, potentially resulting in worse outcome[2,3]. Mechanical circulatory support (MCS) devices, including intra-aortic balloon pump (IABP), microaxial flow pump, and venoarterial extracorporeal membrane oxygenation (VA-ECMO), have been used to mitigate such risks during PCI procedures[4,5]. Unfortunately, randomized trials evaluating IABP or microaxial flow pump in this setting have not demonstrated a significant reduction in mortality or major adverse cardiovascular events[6,7].

Compared with IABP or microaxial flow pump, VA-ECMO can provide superior hemodynamic support and additional respiratory support[8–10]. With advances in device technology and ultrasound guidance, VA-ECMO can be rapidly deployed at the bedside via femoral access[11,12]. Given these advantages, prophylactic VA-ECMO has been increasingly utilized in patients undergoing high-risk PCI despite the lack of robust evidence supporting its clinical benefit[13–15]. However, peripheral VA-ECMO is associated with adverse physiological effects, such as systemic inflammatory responses and increased left ventricular afterload, as well as clinical complications including bleeding, vascular complications, and lower extremity ischemia[16–19]. These adverse physiological effects and complications have led many ECMO centers to adopt a “standby ECMO” strategy, reserving VA-ECMO for emergency use in the setting of refractory hemodynamic compromise during high-risk PCI[20–22]. In a recent single-center randomized trial involving patients undergoing high-risk PCI[23], only 19.4% of patients in the standby ECMO group required emergency VA-ECMO support, suggesting that routine prophylactic use might not be necessary in most cases. Nevertheless, emergency initiation of MCS during PCI has been associated with an increased risk of mortality compared with prophylactic MCS support[24]. Therefore, the optimal strategy of VA-ECMO use in high-risk PCI remains uncertain.

In this context, we propose a novel standby ECMO strategy in which femoral cannulas are pre-inserted and connected to a primed circuit, with arterial and venous lines clamped, and VA-ECMO is initiated only as needed (Figure 1). Accordingly, we designed the ECMO-READY trial to evaluate the comparative effectiveness of pre-cannulated standby cannulated versus prophylactic ECMO strategies in patients undergoing high-risk PCI.

**Figure 1.**
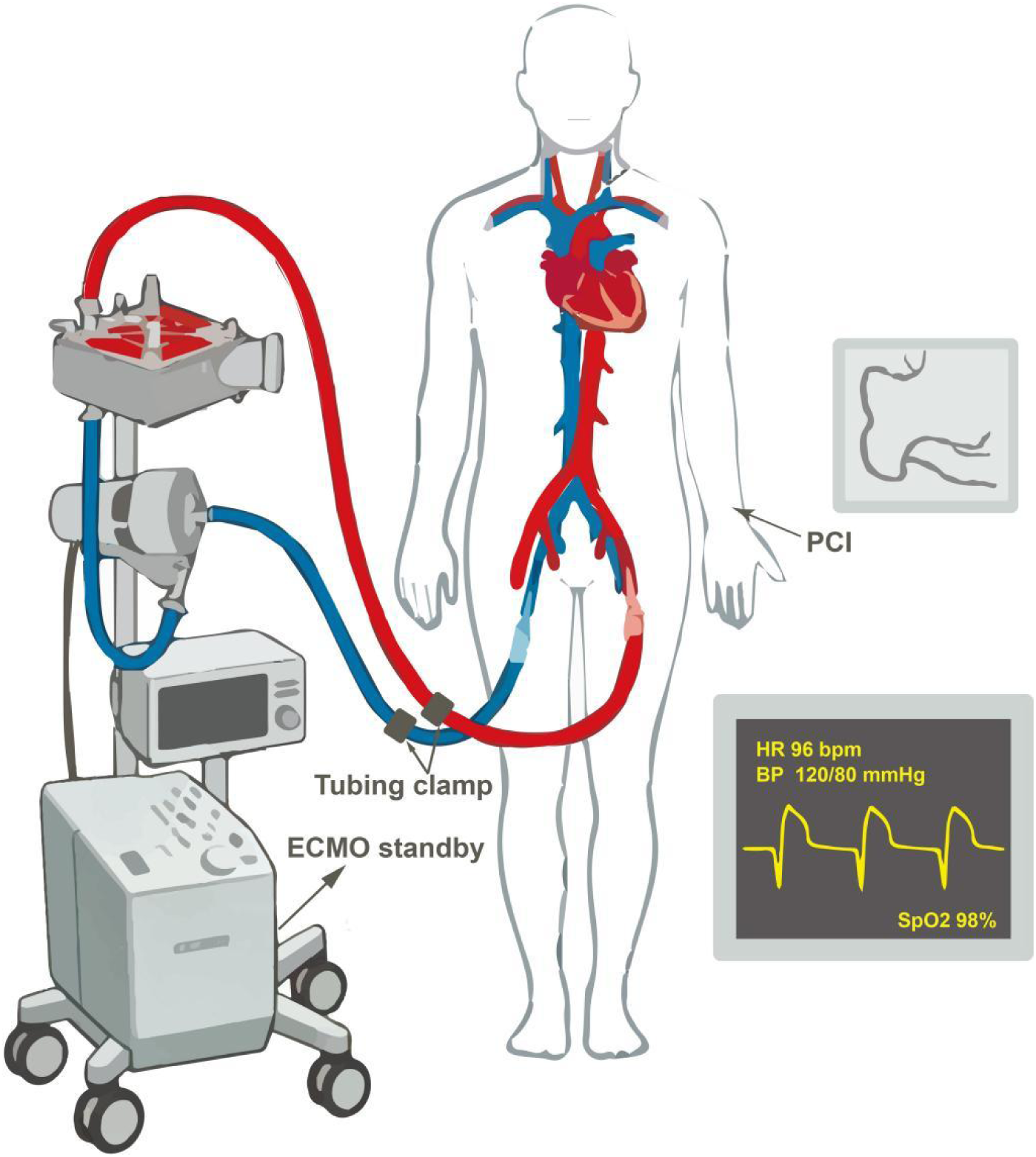
Pre-cannulated standby ECMO strategy. ECMO, extracorporeal membrane oxygenation.

### Objectives {7}

The objective of this trial is to evaluate whether pre-cannulated standby ECMO compared to prophylactic ECMO will reduce the 30-day incidence of major adverse events, including all-cause death, myocardial infraction, any repeat revascularization, stroke, PCI failure, limb ischemia, major bleeding, vascular injury requiring intervention, and need for renal replacement therapy(RRT).

### Trial design {8}

ECMO-READY is a prospective, muticenter, open-label, randomized, parallel-group, superiority trial. Eligible patients who provide written informed consent are randomly assigned in a 1:1 ratio to either the standby ECMO group(n=88) or the prophylactic ECMO group(n=88). The trial design and protocol adhere to the SPIRIT guidelines[25]. Figure 2 illustrates the overall study design.

**Figure 2.**
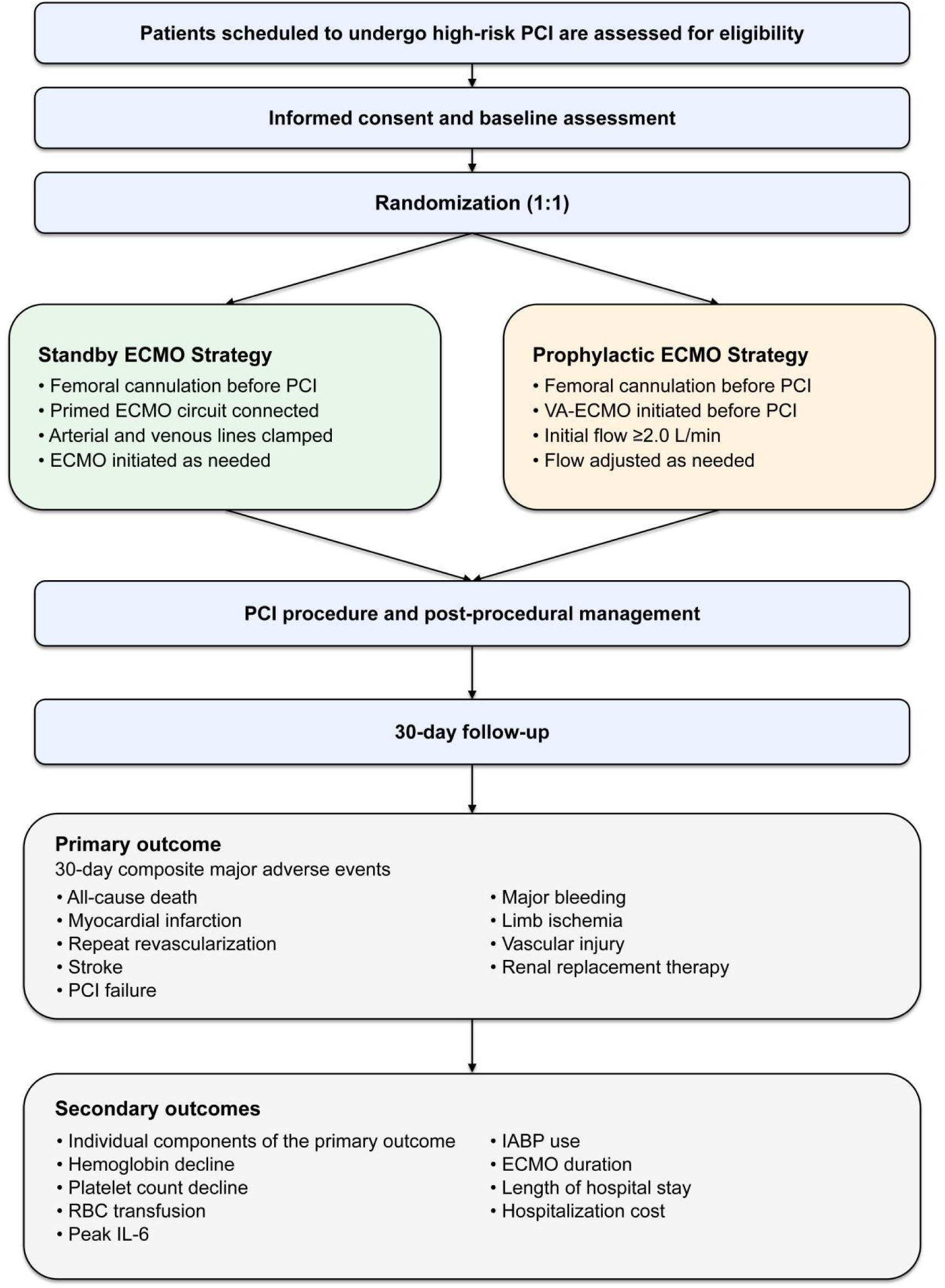
Study design. PCI, percutaneous coronary intervention; ECMO, extracorporeal membrane oxygenation; VA-ECMO, veno-arterial extracorporeal membrane oxygenation; MI, myocardial infarction; RRT, renal replacement therapy; RBC, red blood cell; IL-6, interleukin-6; IABP, intra-aortic balloon pump.

## Methods: Participants, interventions and outcomes

### Study setting {9}

Participants are continuously recruited from the 8 centers in China (see Additional file1: Table S1 for the hospital list).

### Eligibility criteria {10}

Adult patients who are scheduled to undergo high-risk PCI are eligible for enrollment. Inclusion and exclusion criteria are detailed in Table 1.

**Table 1.**
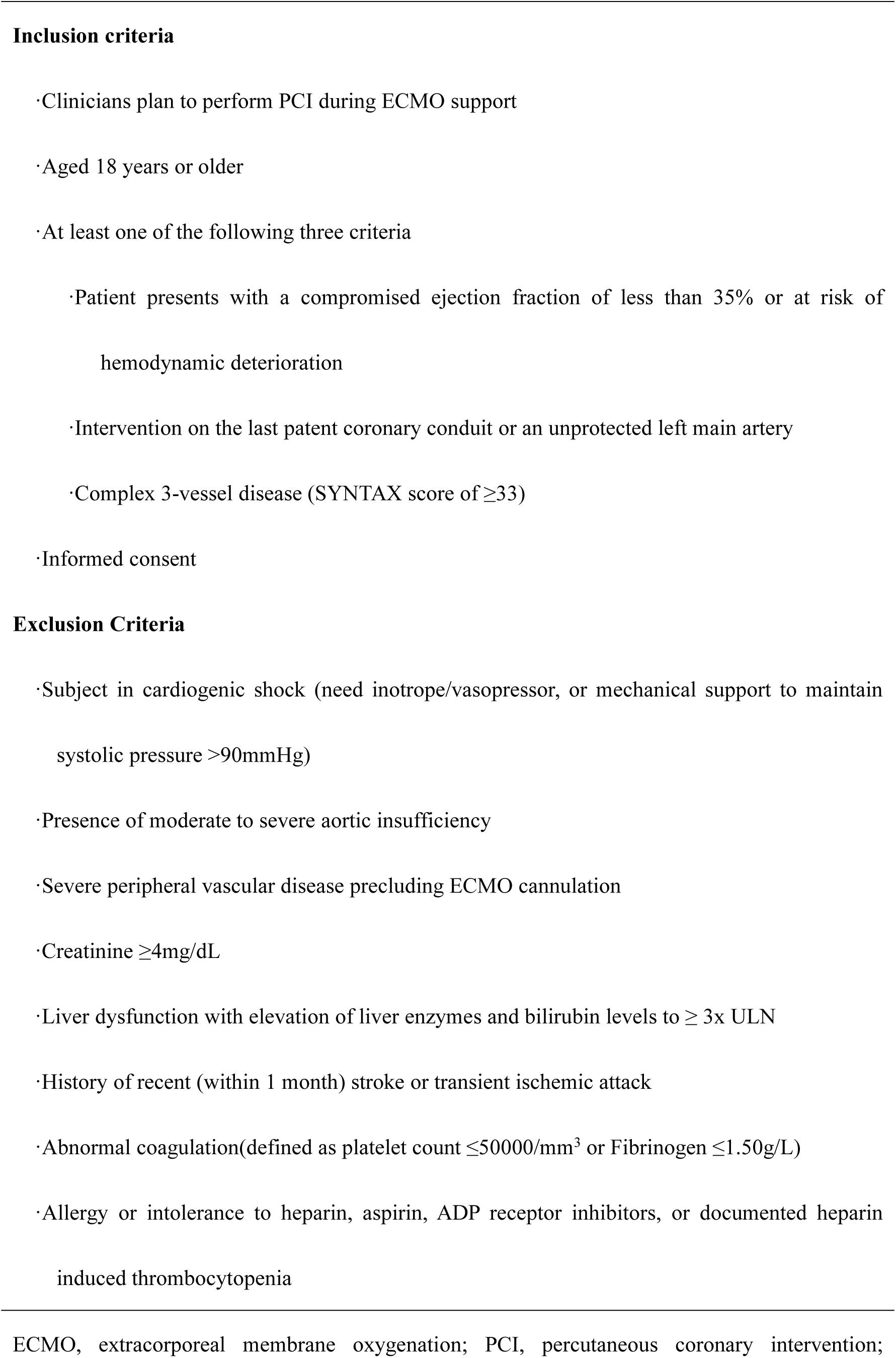
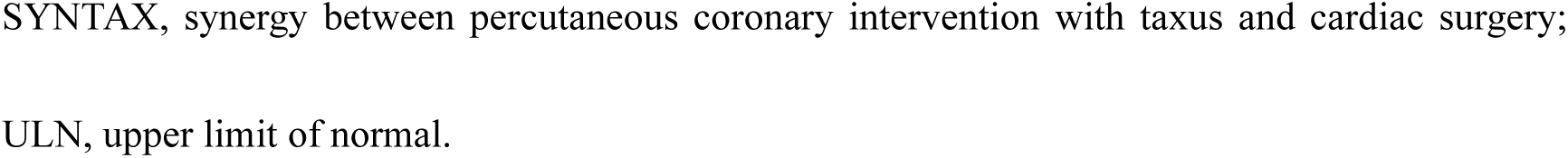
Inclusion and exclusion criteria of the ECMO-READY trial.

### Who will take informed consent? {26a}

Written informed consent will be obtained by the ECMO-READY investigator from each participant prior to enrollment in the clinical trial. During the consent process, the purpose, procedures, potential risks, and benefits of participation will be explained to the patient. If a patient is unable to provide consent due to their clinical condition, written informed consent will be obtained from their legally authorized representative, in accordance with local regulations and institutional policies. Participation will be entirely voluntary, with no coercion.

### Additional consent provisions for collection and use of participant data and biological specimens {26b}

No additional consent is required for the use of study-related clinical data or biological samples.

## Interventions

### Explanation for the choice of comparators {6b}

Prophylactic VA-ECMO was selected as the comparator because it reflects current clinical practice in many centers managing patients undergoing high-risk PCI. However, its routine use remains controversial due to the lack of robust evidence demonstrating clinical benefit and the potential for ECMO-related complications. A pre-cannulated standby ECMO strategy, in contrast, allows for rapid deployment of support only when clinically indicated, potentially reducing unnecessary exposure to ECMO. Therefore, this trial compares prophylactic and standby ECMO strategies to determine the optimal approach to ECMO use in high-risk PCI.

### Intervention description {11a}

In the standby ECMO group, femoral cannulas are inserted via either a percutaneous or surgical approach prior to PCI. The primed ECMO circuit is connected to the cannulas, with arterial and venous lines clamped, and ECMO is maintained in a standby state throughout the procedure. Cannulas are flushed with heparinized saline immediately after insertion to reduce the risk of thrombus formation. In the event of hemodynamic instability precluding completion of the PCI procedure, the clamps are released, and VA-ECMO is promptly initiated to maintain adequate systemic perfusion and hemodynamic support. Femoral cannulas are removed after completion of the PCI procedure.

In the prophylactic ECMO group, femoral cannulas are inserted via either percutaneous or surgical approach prior to PCI. Distal perfusion cannulas are not routinely placed unless clinically indicated (e.g., leg pain, reduced regional oxygen saturation, or skin mottling). The primed ECMO circuit is connected to the cannulas, and VA-ECMO is initiated before the start of PCI. The initial ECMO blood flow is set at 2.0 L/min and subsequently adjusted according to the patient’s mean blood pressure. After completion of PCI, VA-ECMO support is weaned and discontinued at the discretion of the ECMO team, and the femoral cannulas are subsequently removed.

In both groups, 15-17 Fr arterial cannulas and 19-23 Fr venous cannulas are used. All cannulations are performed under ultrasound guidance, with arterial and venous access obtained via the common femoral artery and common femoral vein, respectively. Unfractionated heparin is administered to maintain an activated clotting time of 250-300 seconds during PCI. Arterial cannula removal is performed according to local expertise, with a preference for percutaneous decannulation using interventional closure systems such as suture-based devices.

### Criteria for discontinuing or modifying allocated interventions {11b}

Participants may withdraw from the study at any time, for any reason, without compromising their access to standard medical care. Study discontinuation may also occur in both groups under the following conditions: severe limb ischemia requiring urgent intervention, uncontrolled bleeding, or cannulation-related vascular complications necessitating device revision or removal. Cannulation failure is classified as “intervention not received” rather than study discontinuation. All randomized patients will be followed for outcome assessment regardless of whether the allocated intervention was received, discontinued, or modified.

### Strategies to improve adherence to interventions {11c}

Adherence to the allocated interventions will be promoted through standardized operating procedures, investigator training, and regular study monitoring. Detailed protocols for both standby and prophylactic ECMO strategies, including criteria for ECMO initiation, flow management, anticoagulation, and decannulation, will be provided to all participating centers. Site investigators and study personnel will undergo training prior to study initiation to ensure consistent implementation of the protocol. Monitoring will be conducted at regular 30-day intervals, and will include audits of protocol compliance, data completeness, and measurement accuracy. Feedback will be provided to participating centers to optimize adherence. All audit findings will be documented in a secure, time-stamped audit trail to ensure traceability and support data integrity throughout the study.

### Relevant concomitant care permitted or prohibited during the trial {11d}

All participants will receive guideline-directed medical therapy and standard peri-procedural care for high-risk PCI according to local practice and contemporary recommendations. Permitted concomitant treatments include antiplatelet therapy, anticoagulation, vasopressors, inotropes, mechanical ventilation, RRT, temporary pacing, and other supportive measures deemed clinically necessary by the treating team. Use of additional mechanical circulatory support with an intra-aortic balloon pump after randomization is discouraged unless clinically indicated for left ventricular overload, persistent hemodynamic instability, or ongoing myocardial ischemia. Non-study interventions that could materially influence the primary outcome should be avoided whenever possible. All concomitant treatments and major procedural decisions will be recorded throughout the study period.

### Provisions for post-trial care {30}

After completion of the trial intervention, all participants will continue to receive standard post-procedural and cardiovascular care according to local clinical practice and contemporary guidelines. Management of complications related to PCI, ECMO support, or hospitalization will be provided by the treating institutions. Participants who experience adverse events or complications directly related to trial participation will receive appropriate medical evaluation and treatment in accordance with institutional policies and applicable regulations. Insurance coverage or compensation for trial-related injury, where required, will be provided in accordance with local regulatory and institutional requirements.

### Outcomes {12}

The primary outcome of this trial is the composite rate of intra- and postprocedural major adverse events (MAEs) within 30 days, including all-cause death, myocardial infarction, repeat revascularization, stroke, PCI failure, limb ischemia, major bleeding, vascular injury requiring intervention, and need for RRT. Secondary outcomes include post-procedural hemoglobin decline, post-procedural platelet count decline, red blood cell (RBC) transfusion rate, peak post-procedural interleukin-6 (IL-6) level, use of IABP, duration of ECMO support, length of hospital stay, hospitalization cost, each component of the composite primary outcome. Definitions of all outcomes are provided in the online-only Supplementary Material.

### Participant timeline {13}

The schedule of enrollment, interventions, and assessments is presented in Table 2.

**Table 2.**
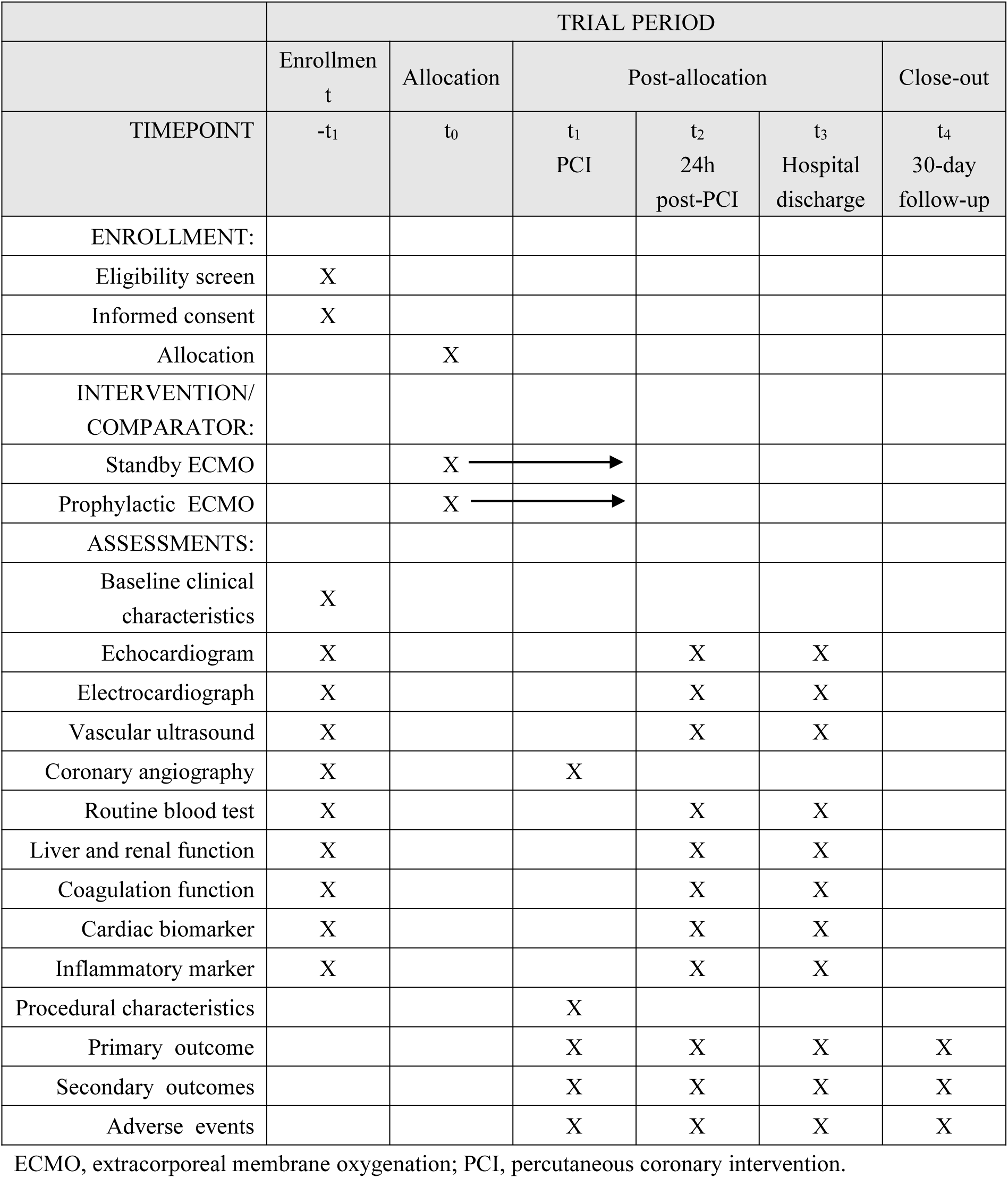
Schedule of enrollment, interventions, and assessments.

|  | TRIAL PERIOD |  |  |  |  |  |
| --- | --- | --- | --- | --- | --- | --- |
|  | Enrollmen<br>t | Allocation | Post-allocation |  |  | Close-out |
| TIMEPOINT | -t<sub>1</sub> | t<sub>0</sub> | t<sub>1</sub><br>PCI | t<sub>2</sub><br>24h<br>post-PCI | t<sub>3</sub><br>Hospital<br>discharge | t<sub>4</sub><br>30-day<br>follow-up |
| ENROLLMENT: |  |  |  |  |  |  |
| Eligibility screen | X |  |  |  |  |  |
| Informed consent | X |  |  |  |  |  |
| Allocation |  | X |  |  |  |  |
| INTERVENTION/<br>COMPARATOR: |  |  |  |  |  |  |
| Standby ECMO |  | X | → |  |  |  |
| Prophylactic ECMO |  | X | → |  |  |  |
| ASSESSMENTS: |  |  |  |  |  |  |
| Baseline clinical<br>characteristics | X |  |  |  |  |  |
| Echocardiogram | X |  |  | X | X |  |
| Electrocardiograph | X |  |  | X | X |  |
| Vascular ultrasound | X |  |  | X | X |  |
| Coronary angiography | X |  | X |  |  |  |
| Routine blood test | X |  |  | X | X |  |
| Liver and renal function | X |  |  | X | X |  |
| Coagulation function | X |  |  | X | X |  |
| Cardiac biomarker | X |  |  | X | X |  |
| Inflammatory marker | X |  |  | X | X |  |
| Procedural characteristics |  |  | X |  |  |  |
| Primary outcome |  |  | X | X | X | X |
| Secondary outcomes |  |  | X | X | X | X |
| Adverse events |  |  | X | X | X | X |

ECMO, extracorporeal membrane oxygenation; PCI, percutaneous coronary intervention.

### Sample size {14}

We based the sample-size calculation on an estimated composite rate of MAEs of 40% in the the prophylactic ECMO group and of 20% in the standby ECMO group using software PASS15.0.5[13, 15]. On the basis of a global type I error of 0.05, we calculated that the enrollment of 158 patients would provide a power of 80% to rule out the null hypothesis of no difference between the two ECMO strategies at a two-sided alpha level of 0.05 for the final analysis. In accordance with an estimated 10% withdrawal, a total of 176 patients will be recruited.

### Recruitment {15}

Potential participants will be identified from patients scheduled to undergo high-risk PCI at participating centers. Screening will be embedded in routine clinical practice through collaboration among interventional cardiology, intensive care, and ECMO teams. Consecutive eligible patients will be approached for enrollment whenever feasible. This multicenter trial will be conducted at 8 high-volume tertiary centers experienced in high-risk PCI and ECMO support. Each site will designate a local investigator and a research coordinator responsible for screening, consent, enrollment, and follow-up. Recruitment progress will be monitored regularly through screening logs and investigator meetings to support timely completion of the target sample size.

## Assignment of interventions: allocation

### Sequence generation {16a}

Participants will be randomly assigned in a 1:1 ratio to the standby ECMO strategy or the prophylactic ECMO strategy according to a computer-generated allocation sequence using permuted blocks with variable block sizes.

### Concealment mechanism {16b}

The allocation sequence will be concealed through a secure centralized web-based randomization system. Access will be granted only after participant eligibility has been confirmed and informed consent has been obtained. Investigators will not have access to future assignments, the full allocation sequence, or enrollment data from other participating centers.

### Implementation {16c}

The randomization sequence will be generated by an independent statistician not involved in participant recruitment or clinical care. Eligible participants will be enrolled by local investigators or research coordinators at each participating center, and treatment allocation will then be assigned automatically through the web-based system.

## Assignment of interventions: Blinding

### Who will be blinded {17a}

Due to the nature of the interventions, this is an open-label trial. Participants, treating clinicians, and site investigators will not be blinded to treatment allocation. Outcome assessors and statisticians will be blinded to treatment allocation whenever feasible.

### Procedure for unblinding if needed {17b}

As this is an open-label trial, formal unblinding procedures are not applicable. All treatment allocations are known to the treating teams at the time of randomization.

## Data collection and management

### Plans for assessment and collection of outcomes {18a}

Baseline, procedural, post-procedural, discharge, and 30-day follow-up data will be collected prospectively using standardized electronic case report forms (eCRFs) at each participating center. Baseline assessments will be performed before randomization and will include demographic and clinical characteristics, echocardiography, electrocardiography, vascular ultrasound, coronary angiography, routine blood tests, liver and renal function, coagulation function, cardiac biomarkers, and inflammatory markers. Procedural data will be collected during the PCI procedure, including PCI characteristics, ECMO parameters, and intra-procedural management. Follow-up assessments will be conducted at 24 hours after PCI, at hospital discharge, and at 30 days after PCI. Echocardiography, electrocardiography, vascular ultrasound, and laboratory tests will be reassessed at 24 hours after PCI and at hospital discharge. Primary and secondary outcomes will be assessed during PCI, at 24 hours after PCI, at hospital discharge, and at 30-day follow-up. Outcome adjudication will be performed independently by assessors blinded to treatment allocation.

To ensure data quality, all study personnel will receive standardized training before study initiation. Data entry will be performed using a secure electronic data capture (EDC) system with built-in range and logic checks. Regular data monitoring and validation will be conducted, and queries will be issued to participating centers to resolve missing or inconsistent data. All data collection procedures will be carried out in accordance with predefined standard operating procedures.

### Plans to promote participant retention and complete follow-up {18b}

Participants will be followed up to 30 days after PCI through scheduled outpatient visits or telephone contact. To promote retention and complete follow-up, contact information will be obtained at enrollment, and reminder calls will be made prior to follow-up assessments. If participants are unable to attend in-person visits, follow-up data will be obtained via telephone contact or review of medical records whenever possible.

All randomized participants will be followed up regardless of adherence to the assigned intervention. Participants who discontinue or deviate from the assigned intervention but consent to continued data collection will remain in follow-up, and primary and secondary outcome data will be collected through medical records, telephone contact, or other available sources. Participants who withdraw consent for data collection will not be contacted further, and no additional data will be collected. **Data management {19}**

All study data will be collected and managed using a secure EDC system. Data entry will be performed by trained study personnel at each participating center, and all data will be coded using unique study identification numbers. The system will incorporate built-in range and logic checks to ensure data accuracy and completeness. Regular data monitoring and validation will be conducted, and data queries will be issued to resolve missing or inconsistent information. Access to the database will be restricted to authorized personnel. All data will be stored on secure servers with routine backup in accordance with institutional and regulatory requirements.

### Confidentiality {27}

Consent forms containing participants’ identifying information will be securely stored at each study site, with access restricted to authorized personnel. Whenever possible, personal identifiers will be removed, and participants will be identified by a unique study identification number. CRFs will be maintained without any identifying information.

### Plans for collection, laboratory evaluation and storage of biological specimens for genetic or molecular analysis in this trial/future use {33}

Not applicable.

## Statistical methods

### Statistical methods for primary and secondary outcomes {20a}

The primary analysis will be performed according to the intention-to-treat (ITT) principle. Sensitivity analyses will be conducted in the per-protocol and as-treated populations to assess the robustness of the findings. The primary outcome will be compared between groups using the chi-square test, and relative risk with corresponding 95% confidence intervals will be estimated to quantify the effect of the standby ECMO strategy. Kaplan–Meier curves will be constructed to illustrate the cumulative incidence of events during the 30-day follow-up period. Effect sizes for secondary outcomes will be reported as relative risks or Hodges-Lehmann estimators, as appropriate, with corresponding 95% confidence intervals. A two-sided significance level of 0.05 will be used.

### Interim analyses {21b}

No interim analyses are planned for this trial.

### Methods for additional analyses (e.g. subgroup analyses) {20b}

Predefined subgroup analyses will be performed across clinically relevant subgroups, including age, sex, use of adjunctive atherectomy, coronary anatomy, and left ventricular systolic function. Treatment effects within subgroups will be presented as relative risks with corresponding 95% confidence intervals and displayed using forest plots. If necessary, adjusted analyses of the primary endpoint will be performed using multivariable regression models to account for clinically relevant baseline covariates that may act as potential confounders.

### Methods in analysis to handle protocol non-adherence and any statistical methods to handle missing data {20c}

No imputation of missing outcome data is planned for the primary analysis. Analyses will be based on available outcome data. If missing outcome data occur, sensitivity analyses will be performed using best-case and worst-case assumptions to assess the robustness of the primary findings. In adjusted analyses, missing data for covariates will be handled using multiple imputation methods.

### Plans to give access to the full protocol, participant level-data and statistical code **{31c}**

The full trial protocol will be submitted as supplementary material alongside the main publication. De-identified participant-level data and statistical code may be shared with qualified researchers upon reasonable request, subject to approval by the study investigators and in accordance with applicable regulations. Data sharing will be conducted in accordance with institutional policies and relevant data protection requirements.

## Oversight and monitoring

### Composition of the coordinating centre and trial steering committee {5d}

The coordinating center is based at Beijing Anzhen Hospital and is responsible for site coordination, investigator communication, recruitment monitoring, data management, and overall trial administration. The trial steering committee, chaired by the principal investigator and composed of study co-investigators, will meet approximately every 3 months or more frequently as needed. The steering committee is responsible for the scientific content of the protocol, oversees the trial operations, and will perform the preparation of the primary manuscript and other publications arising from the ECMO-READY trial. In addition, an independent clinical events committee (CEC), blinded to treatment allocation whenever feasible, will adjudicate all clinical endpoints according to prespecified endpoint definitions.

### Composition of the data monitoring committee, its role and reporting structure **{21a}**

An independent data safety monitoring board (DSMB), composed of experts in interventional cardiology, intensive care, ECMO management, and statistics, has been established. The DSMB is independent of the sponsor and has no competing interests related to the trial. Its primary responsibility is to periodically monitor trial progress and participant safety, and to recommend continuation, modification, or termination of the trial as appropriate.

### Adverse event reporting and harms {22}

All adverse events (AEs) and serious adverse events (SAEs) will be monitored and recorded throughout the study period. AEs may be identified through routine clinical assessments, laboratory testing, review of medical records, or spontaneous reporting by participants or treating clinicians. Site investigators will assess the severity and potential relationship of adverse events to the study intervention. SAEs, including death, cardiac arrest, myocardial infarction, stroke, limb ischemia, major bleeding, vascular injury requiring intervention, and RRT, will be reported to the coordinating center, DSMB, and institutional ethics committees in accordance with local regulatory requirements. All safety reports will be documented in the study database and periodically reviewed by the DSMB. Participants experiencing AEs will receive appropriate management according to standard clinical practice.

### Frequency and plans for auditing trial conduct {23}

The coordinating center and project management team will review recruitment status, protocol adherence, and data quality on a monthly basis. The DSMB will review participant safety and overall study conduct approximately every 3 months, or more frequently if necessary.

### Plans for communicating important protocol amendments to relevant parties (e.g. trial participants, ethical committees) {25}

All important protocol modifications (e.g., changes to eligibility criteria, outcomes, and analyses) will be submitted to the relevant ethics committees for review and approval. Approved protocol amendments will be communicated promptly to investigators, participating centers, trial registries, and other relevant regulatory authorities as required. Trial participants will be informed of protocol changes when relevant to their participation, safety, or informed consent.

### Dissemination plans {31a}

Trial results will be presented at scientific conferences, and published in peer-reviewed journals. De-identified individual participant data will be made available to to qualified researchers upon reasonable request after trial completion.

## Discussion

The optimal strategy for ECMO support in patients undergoing high-risk PCI remains controversial. Although prophylactic VA-ECMO may provide hemodynamic stability during complex PCI procedures[26], it is also associated with potential complications and increased resource utilization[27, 28]. In contemporary clinical practice, some centers have adopted a standby ECMO strategy in which vascular access and ECMO circuits are prepared before PCI, with ECMO initiated only when clinically necessary. However, prospective randomized evidence comparing prophylactic and standby ECMO strategies is limited.

To the best of our knowledge, the ECMO-READY trial is the first randomized trial to evaluate whether a pre-cannulated standby ECMO strategy can reduce the 30-day incidence of MAEs compared with routine prophylactic ECMO in patients undergoing high-risk PCI. One recently published randomized controlled trial compared prophylactic VA-ECMO with a conventional standby ECMO strategy and reported a lower rate of life-threatening complications in the prophylactic ECMO group[23]. However, interpretation of these findings is limited by the small sample size, single-center design, and the definition of life-threatening complications, which primarily reflected intraprocedural hemodynamic instability requiring urgent ECMO initiation in the standby group.

The present trial has several strengths, including its prospective multicenter randomized design, clinically relevant intervention strategy, clinically meaningful primary outcome, and enrollment of patients from experienced cardiovascular centers. In addition, blinded adjudication of clinical endpoints will be performed whenever feasible to reduce assessment bias.

Nonetheless, some limitations and operational considerations should be acknowledged. First, the trial employs an open-label design due to the nature of the intervention, which may introduce performance bias. Second, variations in PCI techniques, ECMO management, and peri-procedural care across participating centers may influence clinical outcomes despite standardized study protocols and investigator training. Third, the decision to initiate emergent ECMO support in the standby group may be influenced by inter-operator and inter-center variability in the assessment of intraprocedural hemodynamic instability. Finally, randomization is not stratified by participating center, which may result in residual center-level heterogeneity.

In conclusion, the ECMO-READY trial is expected to provide important prospective randomized evidence regarding ECMO support strategies for patients undergoing high-risk PCI and may help inform future clinical practice.

## Trial status

The current protocol is version 1.0, dated January 1, 2024. Patient recruitment began in March 2024 and is anticipated to be completed by December 2026.

## Data Availability

All data produced in the present study are available upon reasonable request to the authors

## Abbreviations

ECMO: extracorporeal membrane oxygenation
VA-ECMO: venoarterial extracorporeal membrane oxygenation
PCI: percutaneous coronary intervention
MCS: mechanical circulatory support
IABP: intra-aortic balloon pump
RRT: renal replacement therapy
RBC: red blood cell
IL-6: interleukin-6
eCRF: electronic case report form
EDC: electronic data capture
ITT: intention-to-treat
CEC: clinical events committee
DSMB: data safety monitoring board
AE: adverse event
SAE: serious adverse event
MAE: major adverse event
ULN: upper limit of normal
SYNTAX: synergy between percutaneous coronary intervention with taxus and cardiac surgery
BARC: bleeding academic research consortium

## Supplementary Material

**Additional file 1:** Definitions of study outcomes and participating centers in the ECMO-READY trial (Table S1).

## Acknowledgements

We would like to thank all the participant centers in the ECMO-READY trial.

## Declarations

### Authors’ contributions {31b}

XH is the chief investigator and conceived the study, and led the proposal and protocol development. LW, AL, KW, CL, FL, XY, DW, LL, WD, XW, ZD, HW, YL, and JL contributed to the study design and development of the proposal. LW and EF served as the lead trial methodologists. LW, YW, and AL drafted and revised the manuscript. All authors reviewed and approved the final manuscript.

### Availability of data and materials {29}

The datasets generated and analyzed during the current study will be available from the principal investigator upon reasonable request and subject to approval by the relevant ethics committee.

### Ethics approval and consent to participate {24}

This study protocol was reviewed and approved by the Ethics Committee of Beijing Anzhen Hospital, Capital Medical University (Approval ID: KS2024014). Written informed consent will be obtained from all participants or, when patients are unable to provide consent due to their medical condition, from a legally authorized representative in accordance with institutional policies and applicable national regulations.

### Consent for publication {32}

Not applicable.

## Supplemental digital content

### Definitions of outcomes

#### Components of composite primary outcome

- All-cause death is defined death from any cause occurring from the day of percutaneous coronary intervention (PCI) through 30 days thereafter, regardless of the underlying cause.
- Myocardial infraction is defined according to the the Fourth Universal Definition of Myocardial Infarction. The diagnosis is made when there is acute myocardial injury with clinical evidence of acute myocardial ischaemia and with detection of a rise and/or fall of cardiac biomarkers with at least one value above the 99th percentile URL, together with at least one of the following: (i) symptoms of myocardial ischaemia; (ii) new ST-elevation ≥1 mm, new pathological Q waves, or new left bundle branch block; (iii) imaging evidence of new loss of viable myocardium or new regional wall motion abnormality; (iv) identification of a coronary thrombus by angiography. Peri-PCI myocardial infarction is defined by an elevation of biomarkers > 5 times of the 99th percentile URL.
- Repeat revascularization was defined as the need for repeat PCI or coronary artery bypass grafting.
- Stroke is defined as a newly occurring cerebral infarction or intracranial hemorrhage confirmed by cerebral computed tomography and an attending neurologic consultant, with or without accompanying neurological symptoms or signs(e.g., motor or sensory deficits, altered consciousness, aphasia, or impaired language comprehension).
- PCI failure is defined as inability to complete the intended revascularization of the target lesion with an acceptable angiographic result, including residual stenosis >20% after stent implantation, residual stenosis >50% after balloon angioplasty, or final TIMI flow grade <3 in the target vessel.
- Limb ischemia is defined as lower-extremity ischemia requiring placement of a distal perfusion catheter or surgical/interventional treatment (e.g., fasciotomy, amputation, or endovascular intervention)
- Major bleeding is defined as Bleeding Academic Research Consortium (BARC) type 3 to 5 bleeding.
- Vascular injury requiring intervention is defined as femoral cannulation-site vascular complications(e.g., pseudoaneurysm, arteriovenous fistula, arterial or venous laceration, or dissection) requiring surgical repair or endovascular treatment, regardless of whether the procedure was ultimately performed.
- Need for renal replacement therapy (RRT) is defined as new initiation of renal replacement therapy after randomization because of acute kidney injury, refractory fluid overload, severe electrolyte disturbance, or metabolic acidosis, in patients not receiving renal replacement therapy at baseline.

#### Secondary outcomes

- Post-procedural hemoglobin decline is defined as the relative decrease in hemoglobin from the pre-procedural value to the nadir post-procedural value, calculated as (pre-procedural hemoglobin - nadir post-procedural hemoglobin) / pre-procedural hemoglobin × 100%.
- Post-procedural platelet count decline is defined as the relative decrease in platelet count from the pre-procedural value to the nadir post-procedural value, calculated as (pre-procedural platelet count - nadir post-procedural platelet count) / pre-procedural platelet count × 100%.
- Red blood cell (RBC) transfusion rate is defined as the percentage of patients who receive at least one unit of packed RBCs from randomization to hospital discharge.
- Peak post-procedural interleukin-6 (IL-6) level is defined as the highest serum IL-6 concentration measured after the procedure during hospitalization.
- Use of intra-aortic balloon pump (IABP) is defined as unplanned initiation of IABP after randomization.
- Duration of extracorporeal membrane oxygenation (ECMO) support is defined as the time from initiation of ECMO support to successful decannulation.
- Length of hospital stay is defined as the number of days from PCI to hospital discharge
- Hospitalization cost is defined as the total cost until the discharge from the hospital.
- Each component of the composite primary outcome includes the incidence of all-cause death, myocardial infraction, repeat revascularization, stroke, PCI failure, limb ischemia, major bleeding, vascular injury requiring intervention, or need for RRT.

---

**Table S1.**
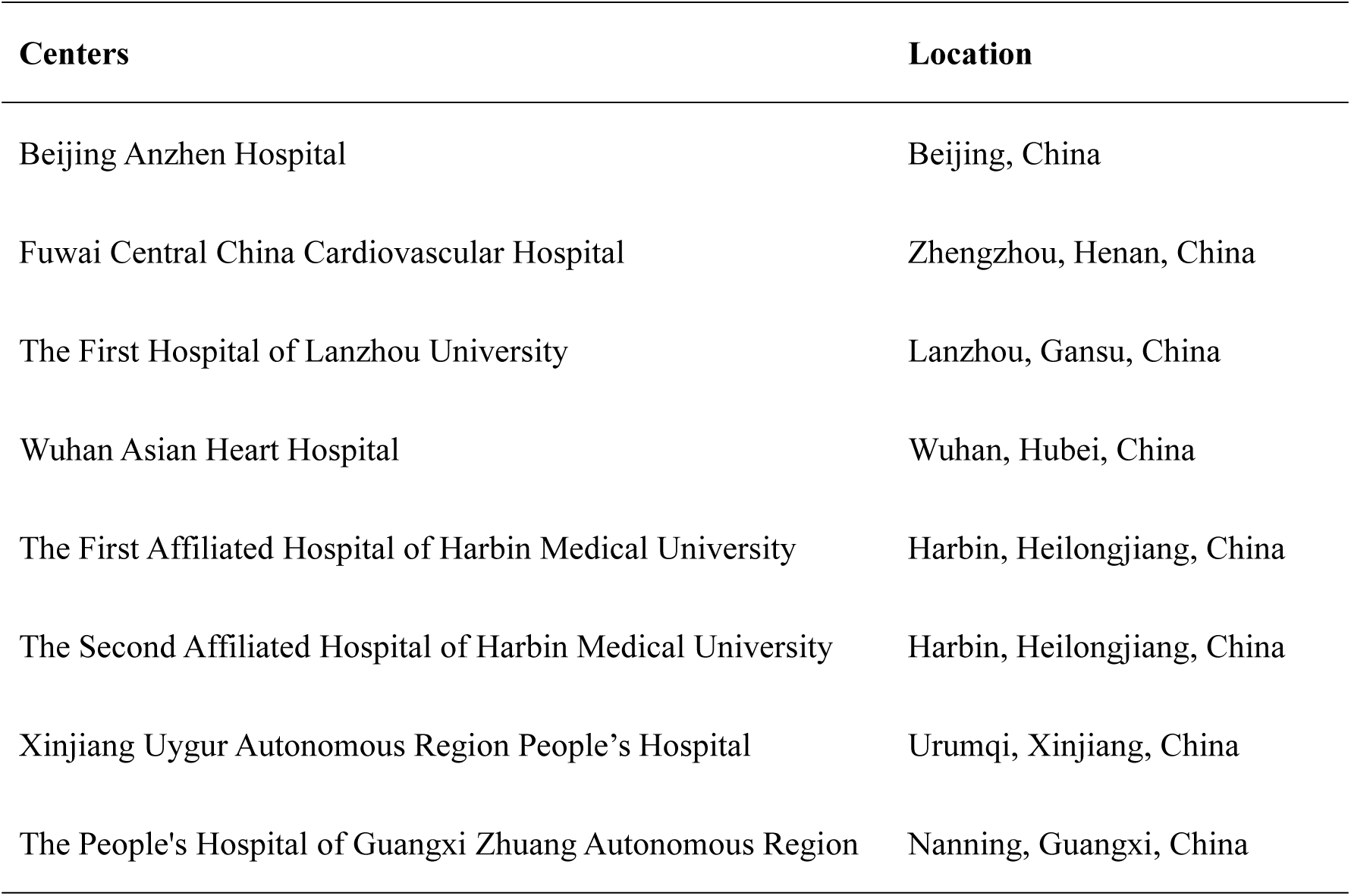
ECMO-READY trial participating centers.

| **Centers** | **Location** |
| --- | --- |
| Beijing Anzhen Hospital | Beijing, China |
| Fuwai Central China Cardiovascular Hospital | Zhengzhou, Henan, China |
| The First Hospital of Lanzhou University | Lanzhou, Gansu, China |
| Wuhan Asian Heart Hospital | Wuhan, Hubei, China |
| The First Affiliated Hospital of Harbin Medical University | Harbin, Heilongjiang, China |
| The Second Affiliated Hospital of Harbin Medical University | Harbin, Heilongjiang, China |
| Xinjiang Uygur Autonomous Region People's Hospital | Urumqi, Xinjiang, China |
| The People's Hospital of Guangxi Zhuang Autonomous Region | Nanning, Guangxi, China |

## References

1. Bricker RS, Glorioso TJ, Jawaid O, Plomondon ME, Valle JA, Armstrong EJ, et al. Temporal Trends and Site Variation in High-Risk Coronary Intervention and the Use of Mechanical Circulatory Support: Insights From the Veterans Affairs Clinical Assessment Reporting and Tracking (CART) Program. J Am Heart Assoc. 2019;8(24):e014906.

2. Mamas MA, Anderson SG, O’Kane PD, Keavney B, Nolan J, Oldroyd KG, et al. Impact of left ventricular function in relation to procedural outcomes following percutaneous coronary intervention: insights from the British Cardiovascular Intervention Society. Eur Heart J. 2014;35(43):3004–12a.

3. Lopes RD, Alexander KP, Stevens SR, Reynolds HR, Stone GW, Piña IL, et al. Initial Invasive Versus Conservative Management of Stable Ischemic Heart Disease in Patients With a History of Heart Failure or Left Ventricular Dysfunction: Insights From the ISCHEMIA Trial. Circulation. 2020;142(18):1725–35.

4. Rihal CS, Naidu SS, Givertz MM, Szeto WY, Burke JA, Kapur NK, et al. 2015 SCAI/ACC/HFSA/STS Clinical Expert Consensus Statement on the Use of Percutaneous Mechanical Circulatory Support Devices in Cardiovascular Care: Endorsed by the American Heart Assocation, the Cardiological Society of India, and Sociedad Latino Americana de Cardiologia Intervencion; Affirmation of Value by the Canadian Association of Interventional Cardiology-Association Canadienne de Cardiologie d’intervention. J Am Coll Cardiol. 2015;65(19):e7–e26.

5. O’Neill WW, Kleiman NS, Moses J, Henriques JP, Dixon S, Massaro J, et al. A prospective, randomized clinical trial of hemodynamic support with Impella 2.5 versus intra-aortic balloon pump in patients undergoing high-risk percutaneous coronary intervention: the PROTECT II study. Circulation. 2012;126(14):1717–27.

6. Perera D, Ryan M, Ezad SM, Khan SQ, Webb I, O’Kane PD, et al. Left Ventricular Unloading in High-Risk Percutaneous Coronary Intervention. N Engl J Med. 2026;394(18):1779–1789.

7. Perera D, Stables R, Thomas M, Booth J, Pitt M, Blackman D, et al. Elective intra-aortic balloon counterpulsation during high-risk percutaneous coronary intervention: a randomized controlled trial. JAMA. 2010;304(8):867–74.

8. Grand J, Udesen NLJ, Bro-Jeppesen J. Mechanical circulatory support after cardiac arrest. Curr Opin Crit Care. 2025;31(6):717–22.

9. Moller JE, Thiele H, Zeymer U, Proudfoot A, Hassager C. Mechanical circulatory support for patients with infarct-related cardiogenic shock: a state-of-the-art review. Heart. 2025;111(10):445–53.

10. Thiele H, Zeymer U, Akin I, Behnes M, Rassaf T, Mahabadi AA, et al. Extracorporeal Life Support in Infarct-Related Cardiogenic Shock. N Engl J Med. 2023;389(14):1286–97.

11. Wang L, Li C, Hao X, Rycus P, Tonna JE, Alexander P, et al. Percutaneous cannulation is associated with lower rate of severe neurological complication in femoro-femoral ECPR: results from the Extracorporeal Life Support Organization Registry. Ann Intensive Care. 2023;13(1):77.

12. Wang L, Yang F, Zhang S, Li C, Du Z, Rycus P, et al. Percutaneous versus surgical cannulation for femoro-femoral VA-ECMO in patients with cardiogenic shock: Results from the Extracorporeal Life Support Organization Registry. J Heart Lung Transplant. 2022;41(4):470–81.

13. Cheng A, Wang B, Fu G, Li C, Liu Y, Li F, et al. Long-Term Outcomes of Patients Undergoing Extracorporeal Membrane Oxygenator-Assisted High-Risk Percutaneous Coronary Intervention. Catheter Cardiovasc Interv. 2025;106(4):2637–44.

14. Zuin M, Rigatelli G, Daggubati R. Cardiac intensive care management of high-risk percutaneous coronary intervention using the venoarterial ECMO support. Heart Fail Rev. 2020;25(5):833–46.

15. van den Brink FS, Meijers TA, Hofma SH, van Boven AJ, Nap A, Vonk A, et al. Prophylactic veno-arterial extracorporeal membrane oxygenation in patients undergoing high-risk percutaneous coronary intervention. Neth Heart J. 2020;28(3):139–44.

16. Yang F, Hou D, Wang J, Cui Y, Wang X, Xing Z, et al. Vascular complications in adult postcardiotomy cardiogenic shock patients receiving venoarterial extracorporeal membrane oxygenation. Ann Intensive Care. 2018;8(1):72.

17. Wang K, Wang L, Ma J, Xie H, Li C, Hao X, et al. Intra-aortic balloon pump after VA-ECMO reduces mortality in patients with cardiogenic shock: an analysis of the Chinese extracorporeal life support registry. Crit Care. 2024;28(1):394.

18. Millar JE, Fanning JP, McDonald CI, McAuley DF, Fraser JF. The inflammatory response to extracorporeal membrane oxygenation (ECMO): a review of the pathophysiology. Crit Care. 2016;20(1):387.

19. Nunez JI, Gosling AF, O’Gara B, Kennedy KF, Rycus P, Abrams D, et al. Bleeding and thrombotic events in adults supported with venovenous extracorporeal membrane oxygenation: an ELSO registry analysis. Intensive Care Med. 2022;48(2):213–24.

20. Wang L, Wang Y, Fu H, Meng X, Li C, Jiang C, et al. Feasibility of standby ECMO with preset femoral vascular sheaths for high-risk transcatheter aortic valve replacement. Perfusion. 2025;40(4):893–7.

21. Liu C, Li X, Li J, Shen D, Sun Q, Zhao J, et al. Standby extracorporeal membrane oxygenation: a better strategy for high-risk percutaneous coronary intervention. Front Med (Lausanne). 2024;11:1404479.

22. Teirstein PS, Vogel RA, Dorros G, Stertzer SH, Vandormael MG, Smith SC, Jr., et al. Prophylactic versus standby cardiopulmonary support for high risk percutaneous transluminal coronary angioplasty. J Am Coll Cardiol. 1993;21(3):590–6.

23. Pan C, Zhu Y, Zhao J, Zhang B, Hu S, Zhao C, et al. Prophylactic VA-ECMO During Complex High-Risk PCI: A Randomized Controlled Trial. JACC Adv. 2025;4(9):102095.

24. O’Neill BP, Grines C, Moses JW, Ohman EM, Lansky A, Popma J, et al. Outcomes of bailout percutaneous ventricular assist device versus prophylactic strategy in patients undergoing nonemergent percutaneous coronary intervention. Catheter Cardiovasc Interv. 2021;98(4):E501–e12.

25. Chan AW, Tetzlaff JM, Gøtzsche PC, Altman DG, Mann H, Berlin JA, et al. SPIRIT 2013 explanation and elaboration: guidance for protocols of clinical trials. BMJ. 2013;346:e7586.

26. Lüsebrink E, Binzenhöfer L, Hering D, Villegas Sierra L, Schrage B, Scherer C, et al. Scrutinizing the Role of Venoarterial Extracorporeal Membrane Oxygenation: Has Clinical Practice Outpaced the Evidence? Circulation. 2024;149(13):1033–52.

27. Murakami T, Sakakura K, Jinnouchi H, Taniguchi Y, Tsukui T, Watanabe Y, et al. Complications related to veno-arterial extracorporeal membrane oxygenation in patients with acute myocardial infarction: VA-ECMO complications in AMI. J Cardiol. 2022;79(2):170–8.

28. Bogerd M, Ten Hoorn L, Ten Berg S, Peters EJ, Engström AE, Malekzadeh A, et al. Resource utilization associated with extracorporeal membrane oxygenation vs. microaxial flow pump for infarct-related cardiogenic shock. Eur Heart J Acute Cardiovasc Care. 2025;14(5):279–87.

